# In-depth profiling of the immuno-virological landscape after a decade of sustained HIV-1 suppression following ART resumption

**DOI:** 10.1101/2023.11.14.23298452

**Authors:** Cynthia Lungu, Tanvir Hossain, Raquel Crespo, Henrieke A.B. Prins, Kathryn S. Hensley, Casper Rokx, Shringar Rao, Jeroen J. A. van Kampen, David A.M.C. van de Vijver, Thibault Mesplède, Peter D. Katsikis, Rob A. Gruters, Yvonne M. Mueller, Tokameh Mahmoudi

**Affiliations:** Department of Pathology, Erasmus University Medical Center, Rotterdam, Netherlands; Department of Internal Medicine, Section Infectious Diseases, Erasmus University Medical Center, Rotterdam, Netherlands; Department of Medical Microbiology and Infectious Diseases, Erasmus University Medical Center, Rotterdam, Netherlands; Department of Biochemistry, Erasmus University Medical Center, Rotterdam, Netherlands; Department of Viroscience, Erasmus University Medical Center, Rotterdam, Netherlands; Department of Immunology, Erasmus University Medical Center, Rotterdam, Netherlands; Department of Urology, Erasmus University Medical Center, Rotterdam, Netherlands

## Abstract

Despite the success of antiretroviral therapy (ART) in suppressing HIV-1 replication, viral reservoirs persist and reignite viremia upon treatment interruption, posing a major barrier to achieving a cure. Analytical treatment interruption (ATI) is a key experimental strategy in HIV-1 cure research, used to evaluate the efficacy of novel therapeutic interventions. While ATI protocols have evolved to minimize clinical risks, the long-term consequences of ATI remain poorly understood. Here, we investigated the enduring immunologic and virologic effects of extended ATI in a unique cohort of volunteers enrolled in a dendritic cell–based therapeutic vaccine trial (DC-TRN) nearly two decades ago. Participants were re-evaluated more than 10 years after ART resumption. Using integrated approaches—including proviral reservoir quantification, inducibility profiling and high-dimensional immune phenotyping—we identified a significant expansion of the inducible HIV-1 reservoir and a distinct immune signature, despite sustained viral suppression and clinical stability. Notably, Central Memory T cells expressing CXCR3 positively correlated with inducible *tat/rev* msRNA+ T cells. These findings underscore the durable imprint of ATI-based immune interventions on HIV-1 reservoir dynamics and immune homeostasis. Overall, the findings emphasize the need for extended follow-up and functional immune assessments in HIV-1 cure-focused trials.

## INTRODUCTION

Although more than four decades have passed since the discovery of human immunodeficiency virus type 1 (HIV-1), a curative intervention remains elusive. While combination antiretroviral therapy (ART) has transformed HIV-1 into a manageable chronic condition, it does not eradicate the virus. A heterogenous reservoir of latently infected cells (1–3) can persist for decades despite potent ART and reignite systemic viremia upon ART interruption or failure (4, 5). Compounding this challenge is the extreme rarity of latently infected cells, estimated between 1 and 1000 per million CD4⁺ T cells, and the inability of current assays to reliably distinguish between replication-competent and defective proviruses (6–8). Furthermore, tissue reservoirs, particularly within lymphoid, gut-associated, and central nervous system compartments, pose additional barriers due to limited accessibility and distinct immunological microenvironments (9, 10). Consequently, strategies aiming to reduce or eliminate the HIV-1 reservoir must be assessed not only for their biological efficacy but also for their ability to sustain virologic control in the absence of ART. In this context, analytical treatment interruption (ATI), an intentional and closely monitored cessation of ART, has emerged as a critical experimental tool to evaluate the effectiveness of cure-directed interventions (11–13).

ATI protocols have evolved significantly since their initial implementation in the early 2000s (14). The landmark SMART study underscored the potential dangers of antiretroviral treatment pauses, which were associated with increased morbidity and mortality (15). These findings spurred the development of more refined ATI strategies incorporating stringent monitoring, standardized criteria for ART resumption, and ethical oversight (14, 16). Today, ATI remains indispensable in HIV cure research, offering a functional readout of viral rebound kinetics and immune control in response to experimental therapies, including latency-reversing agents, therapeutic vaccines, and broadly neutralizing antibodies (17–19). Despite improved safety measures in current trial designs, ATI carries residual risks associated with uncontrolled viral rebound such as increased transmission and potential heightened immune activation. In addition, and critical to underline, studies have demonstrated that the reservoir is established very early on upon infection (20–22). Within a few weeks after discontinuation of antiretroviral therapy, there is a rapid increase in plasma HIV-1 RNA, a marker of active viral replication. The extent to which differences in the ART-free time after viral rebound (weeks versus months) or the duration and levels of plasma viremia (log 3 versus log 4 or log 5 HIV-1 RNA copies/mL) may enhance HIV-1 reservoir expansion is currently incompletely understood. Therefore, while the stricter guidelines have addressed adverse clinical and immunological consequences of extended periods of viral rebound during ATI (23, 24), they do not address the potential expansion of the HIV-1 reservoir.

Importantly, while short-term immunologic and virologic dynamics during ATI have been well studied (25–30), data on the long-term consequences, particularly after ART-mediated re-suppression are sparse. Recent investigations have begun to address these gaps, with emerging evidence suggesting that ATI may have subtle yet durable impacts on HIV-1 reservoir composition (27, 31). However, comprehensive longitudinal data are still needed to fully understand the implications of ATI and to inform the design of future cure-focused trials.

To build on this emerging evidence and address existing gaps, our study provides an in-depth analysis of the immune-virological landscape following ATI and sustained HIV-1 re-suppression after ART-resumption. We leveraged a rare cohort of individuals who underwent ATI as part of a dendritic cell–based therapeutic vaccine trial (DC-TRN) nearly two decades ago (32). Therapeutic vaccination and subsequent ATI both had a profound and lasting impact on various immune parameters, as evidenced by transcriptome analysis of the blood (33). These individuals were re-enrolled into a follow-up study more than 10 years after ART resumption, which offered an unprecedented opportunity to examine the long-term immunologic and virologic footprint of their past ATI-based immune intervention. By using HIV-1 proviral reservoir quantification, inducibility profiling, and high-dimensional immune phenotyping, we demonstrate a significant expansion of the inducible latent reservoir post-ATI, and characterize the presence of a specific immune signature, despite apparent clinical recovery. These findings offer critical insight into the long-term impact of ATI-based interventions and highlight the value of incorporating extended follow-up and functional immune assessments into future HIV-1 cure trials.

## RESULTS

### Longitudinal Clinical Outcomes Following Prolonged ATI and Resumption of ART

Our study cohort offers a rare opportunity to examine long-term immunovirological outcomes following vaccination and analytical treatment interruption (ATI). Seventeen chronically treated individuals with HIV-1 subtype B underwent prolonged ATI during a non-randomized, phase I/IIa autologous dendritic cell (DC)-based immunotherapy trial starting in 2006 (International Clinical Trial Registry Platform: Netherlands Trial Registry No. NTR2198) (32). At the time of enrollment for the clinical trial, all 17 participants had been on suppressive ART for a median of 4.8 years (interquartile range (IQR): 4.6-5), with undetectable plasma HIV-1 RNA levels (<50 copies/mL) and CD4⁺ T cell counts consistently above 500 cells/μL (32). While the DC vaccine was shown to be safe and well-tolerated, the elicited immune responses were not sufficient to control viral replication in absence of ART. All participants experienced viral rebound, with plasma HIV-1 RNA levels exceeding 1000 copies/mL within two to eight weeks after ATI (32). The median time off ART was 50 weeks (IQR: 28.5-96), after which therapy was resumed according to protocol-defined CD4+ T cell thresholds or earlier treatment guidelines (32). Detailed clinical characteristics and a treatment history of the DCTRN participants before and after ATI are described elsewhere (32). In 2021, more than a decade later, we re-enrolled nine of the DCTRN trial participants, who were in care at Erasmus Medical Center in the Netherlands, for follow-up sampling to enable a unique longitudinal assessment spanning pre- and post-vaccination, ATI phase, and long-term ART re-suppression. The clinical characteristics are summarized in Supplementary Table 1.

Blood samples were collected at six defined time points: baseline (T0, prior to DC vaccination); pre-ATI (T1, one week after the second DC vaccination; and T2, one week after the fourth dose); early ATI (T3, four weeks after ATI initiation); late ATI (T4, >24 weeks into ATI and immediately prior to ART resumption) (32) and post-ATI (T5, >2 years after ART resumption and T6, >10 years after ART resumption) (Fig. 1A). Based on retrospective clinical data, plasma antiretroviral drug levels measured one month after resuming ART were within therapeutic ranges. Plasma viremia re-suppressed to below 50 copies/mL in all individuals within 18 months of ART resumption (Fig. 1B). Over the following decade, three participants experienced isolated viral blips (50–400 copies/mL) (see note under Supplementary Table 1), with no evidence of ART failure, non-adherence, or use of potential HIV-1 reservoir-targeting therapies. A significant decline in CD4+ T cell counts during ATI was observed, particularly at the late interruption point (T4), where median counts dropped markedly compared to baseline (P<0.0001) (Fig. 1C). CD4+ T cell counts gradually recovered after ART resumption, reaching levels comparable to T0 by T6 (>10 years post-ATI; P=0.9668). Similarly, the CD4+/CD8+ T cell ratio decreased during ATI, with a significant reduction at T4 (vs. T0, P = 0.0002) indicating transient immune dysregulation (Fig. 1D). Although this ratio remained significantly lower than baseline two years after ART resumption (T5 vs. T0, P=0.0095), it returned to pre-ATI levels by T6 (P=0.4668), suggesting long-term immune reconstitution. These data suggest that, although ATI induces statistically significant but transient immune disruption, long-term ART-mediated viral re-suppression supports the restoration of systemic immune homeostasis. However, as explored in subsequent sections, these superficial recoveries may mask deeper, persistent virological and immunological remodeling.

**Fig. 1.**
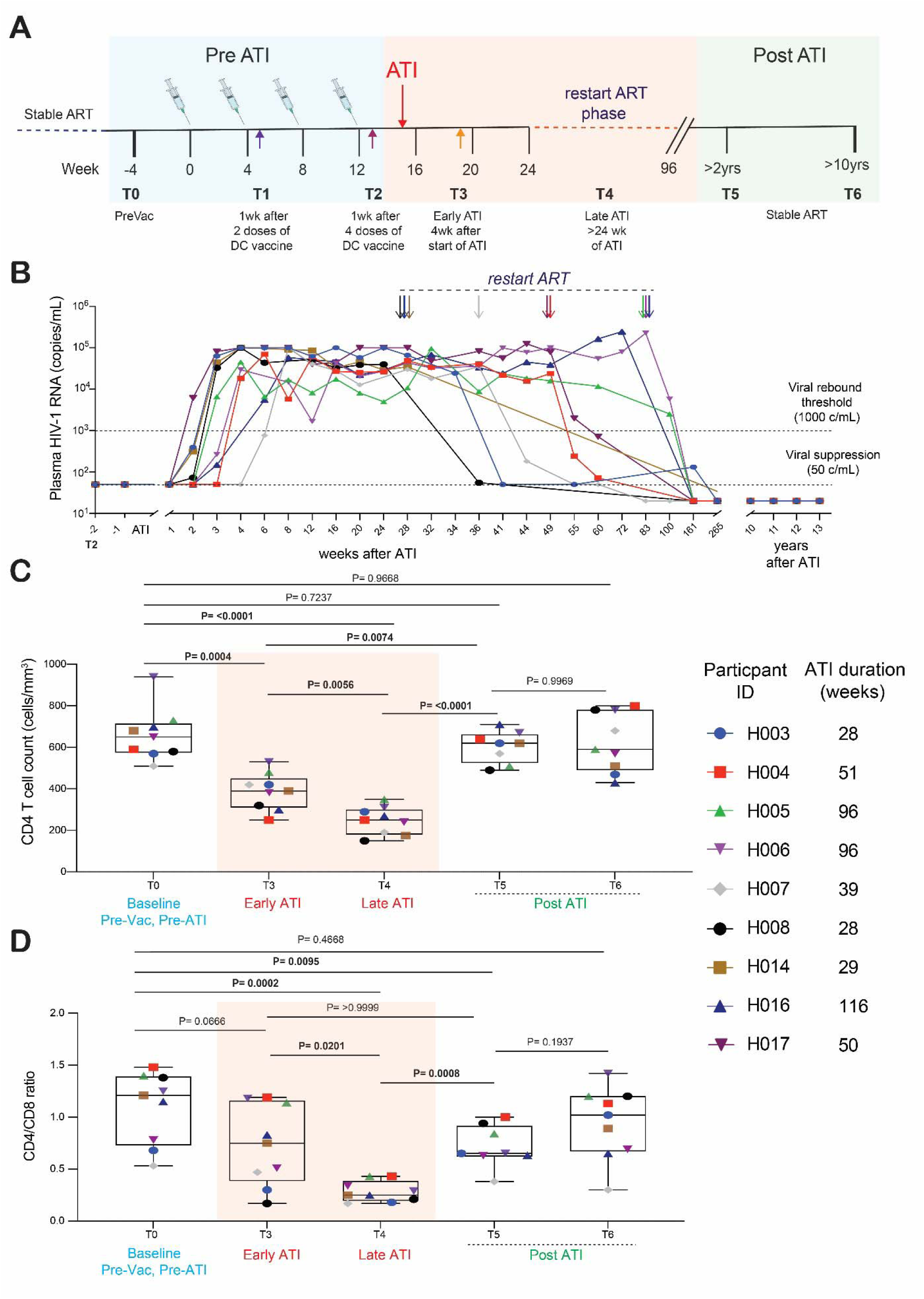
Longitudinal Plasma HIV-1 RNA and T cell dynamics. **A)** Schematic overview of the longitudinal study phase during the DCTRN vaccination trial, ATI (pre-Vac to 96 weeks) and after 13 years on suppressive ART. The study timepoints are indicated as T0 to T6. The start of analytical treatment interruption (ATI) is shown with a red downward arrow. The period during which participants restarted ART is indicated using a dotted horizontal line. Participants received standard care after restarting ART. Timepoint 5 (T5) was a routine care visit with standard blood draw, and no extra samples were collected for reservoir analyses. Timepoint 6 (T6) was an extra visit with large blood collection a median of 12.4 years after resuming ART. **B)** Longitudinal plasma HIV-1 RNA kinetics after ATI. The downward colored arrows mark the time of ART resumption. C) CD4+ T cell counts and D) CD4/CD8 ratio dynamics measured at selected study timepoints. Box plots with mean and range are shown. The shaded bars depict the ART interruption phase. Statistics inferred using a mixed effect analysis with multiple comparisons of the means (Tukey-Kramer test). Adjusted P values are shown above the graphs.

Molecular Dissection of the HIV-1 Reservoir Reveals Long-Term Persistence and Increase of Inducible Latent Reservoir Post-ATI Building upon our longitudinal clinical analyses, we next performed in-depth molecular reservoir quantification to evaluate changes in HIV-1 persistence markers. The extended follow-up and availability of samples at all key phases - pre-ATI, during ATI, and more than a decade after ART resumption - allowed us to comprehensively profile the HIV-1 reservoir across three distinct molecular levels: proviral DNA, viral RNA transcription, and Gag p24 protein translation. This multi-layered assessment provides unique insight into how clinical interventions, including ATI, may influence reservoir composition and stability, helping refine strategies to evaluate the impact of future ATI-based cure interventions.

We first quantified total, defective (5R or 3R deletion), and intact HIV-1 proviral DNA using the Intact Proviral DNA Assay (IPDA) across all key timepoints (Fig. 2A) (7). DC vaccination alone did not alter total proviral DNA levels between T0 and T2 (P=0.9563). However, the proportions of intact proviral DNA increased in two participants, and decreased in four participants. (Supplementary Fig. 1). During ATI, the levels of intact proviral DNA increased at both early (T3) and late (T4) ATI, coinciding with active viremia (Fig. 2A-B). Notably, by T6 (>10 years post-ATI), levels of total, defective, and intact proviral DNA were overall similar to those measured at T0 and T2 (P=0.3533), although four participants showed a notable upward trend in intact proviral DNA compared to T2 (Fig. 2A-B and Supplementary Fig. 1).

**Fig. 2.**
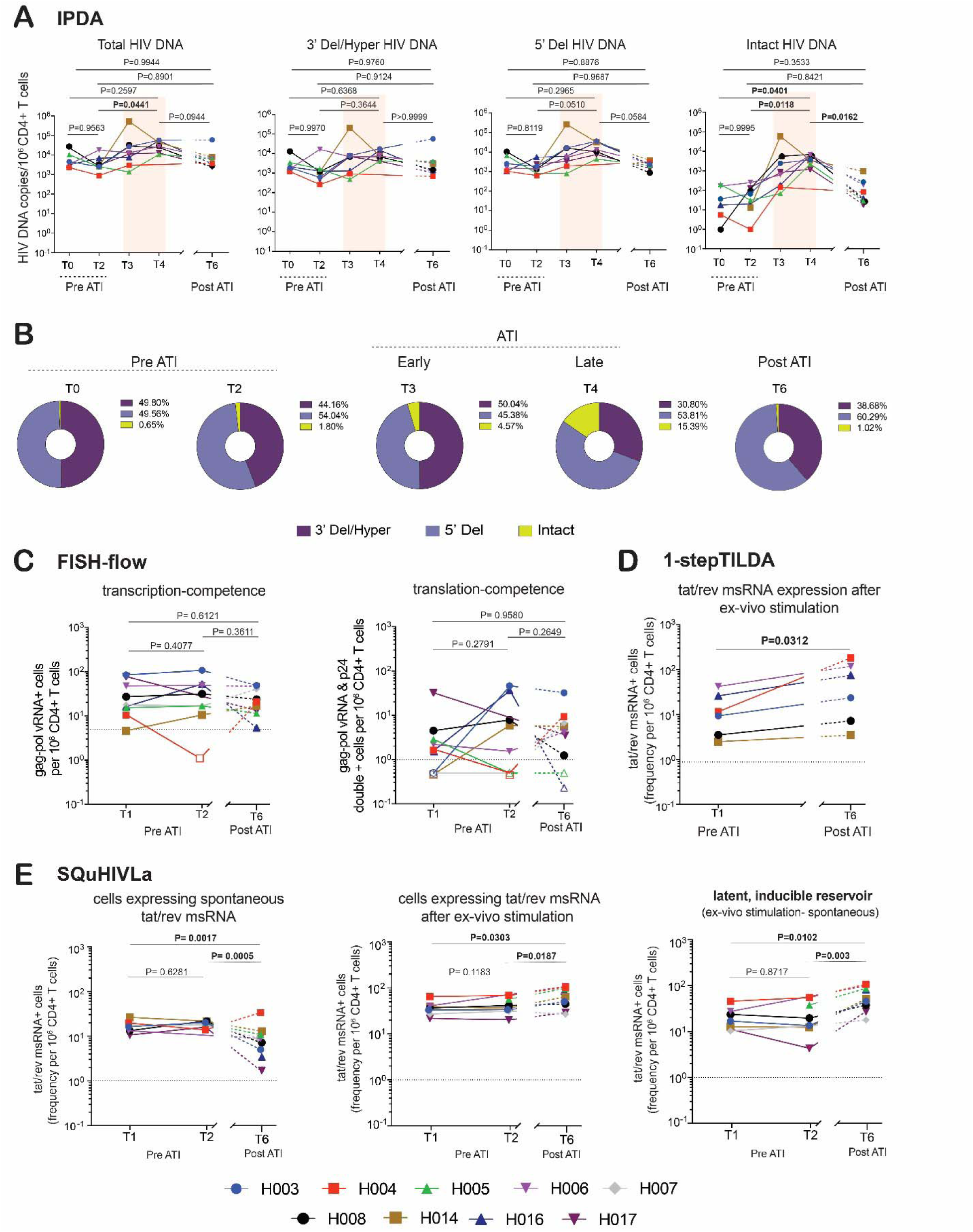
HIV-1 reservoir dynamics after ATI and long-term ART-mediated viral resupression. **A)** Changes in levels of total, intact and defective proviral DNA in peripheral CD4+ T cells in response to DC vaccination, ATI and post-ATI (the shaded bars depict the ATI phase). Mixed effects model analyses and Tukey’s multiple comparison tests were used to compare viral reservoir quantities across timepoints (Adjusted P values are shown). **B)** Change in proportions of intact to defective HIV-1 DNA. **C)** Change in CD4+ T cells expressing unspliced gag-pol viral RNA and cells co-expressing gag-pol vRNA and Gag p24 protein after ex vivo treatment with PMA/ionomycin for 18 hours, measured using FISH-flow. Open symbols depict quantities below the assay limit of detection (dotted line). **D)** Change in CD4+ T cells expressing tat/rev msRNA after ex vivo treatment with PMA/ionomycin for 12 hours, measured using 1-step TILDA. **E)** Change in CD4+ T cells spontaneously expressing tat/rev msRNA (in vivo response) and inducibly expressing tat/rev msRNA after ex vivo treatment with PMA/ionomycin for 12, measured using SQuHIVLa. Mixed effects model analyses and Tukey’s multiple comparison test were used to compare viral reservoir quantities across timepoints. Adjusted P values are shown.

While measurement of the intact viral reservoir at the DNA level by IPDA is considered a marker of replication-competence, the integration site of intact proviruses directly affects inducibility and transcriptional output (34–36). To specifically profile proviral transcriptional activity, we next assessed viral RNA expression using FISH-flow, which detects CD4+ T cells expressing unspliced gag-pol HIV-1 RNA following ex vivo PMA-ionomycin stimulation (37). Median frequencies of gag-pol vRNA+ cells remained stable across T1 (17.85 per 10⁶ CD4+ T cells; IQR: 12.84-62.60), T2 (24.11; IQR: 12.05-51.34), and T6 (17.60; IQR: 12.32-43.62), indicating persistence of the transcription-competent viral reservoir over time (Fig. 2C). Parallel analysis of the translation-competent reservoir, cells co-expressing gag-pol vRNA and Gag p24 protein, revealed consistently low to undetectable frequencies across all timepoints, consistent with previous literature and supporting the notion that viral translation is tightly restricted under suppressive ART (Fig. 2C) (37, 38).

Although HIV-1 RNA expression is universally used as a surrogate for “latency reversal” in cells from participants following ex vivo treatment or after administration of latency reversing agents in vivo (39), most HIV-1 RNA expression comes from defective proviruses. The majority, over 94% of proviruses, is 3’ defective but expresses 3’ defective RNA (40). Tat/rev spliced RNA transcripts are less likely to arise from defective proviruses, and are thus considered a more accurate surrogate marker of replication-competence. Therefore, to more specifically quantify the inducible latent HIV-1 reservoir, we applied two complementary assays targeting tat/rev multiply spliced RNA (msRNA): SQuHIVLa (41) and 1-step TILDA. At T1 and T2, spontaneous tat/rev msRNA expression was comparable between both timepoints (medians: 16.32 and 19.31 per 10⁶ CD4+ T cells, respectively) but declined significantly by T6 (median 8.01; IQR: 3.84-10.38), with statistically significant differences between T6 vs. T1 (P=0.0017) and T6 vs. T2 (P=0.0005), suggesting reduced basal transcriptional activity post-ATI (Fig. 2E). However, upon ex vivo stimulation with PMA-ionomycin, the frequencies of tat/rev msRNA+ cells were significantly higher at T6 than at both T1 and T2 (P=0.0303 and P=0.0187, respectively), indicating increased inducibility of latent proviruses (Fig. 2E). These findings were corroborated by 1-step TILDA, which showed similar trends (P=0.0312) (Fig. 2D). Calculation of the latent, inducible reservoir, defined as the difference between induced and spontaneous tat/rev msRNA+ cells, revealed a statistically significant increase at T6 compared to T1 and T2 (P=0.0102 and P=0.0030, respectively) (Fig. 2F), suggesting an increase of inducible but transcriptionally silent proviruses after more than a decade following ATI.

Since the DC vaccine trial lacked a placebo control, we compared the participants, after more than a decade post-ATI (T6), to a historical cohort of people (n=8) with HIV-1 subtype B matched for key clinical parameters, including sex, age, CD4+ T cell nadir, CD4+ T cell counts at sampling, and years on ART (Supplementary Fig.2 A-D). We leveraged PBMC samples from the historical control participants as a comparator group to assess tat/rev msRNA expression. Using SQuHIVLa, we observed both a significantly higher frequency of CD4+ T cells spontaneously expressing tat/rev msRNA (P=0.0441) as well as significantly higher frequencies of tat/rev msRNA expressing cells after ex vivo PMA-ionomycin stimulation for the post ATI participants at T6 compared to the historical control cohort, (P=0.0437) (Fig. 3A-B). These findings support the notion that ATI may leave a durable imprint on the inducible HIV-1 reservoir, even in individuals who re-suppress viremia and maintain long-term ART. The apparent increase in the inducible HIV-1 reservoir more than a decade post ATI raises important questions about clinical significance and potential role of these cells in future viral rebound events.

**Fig. 3.**
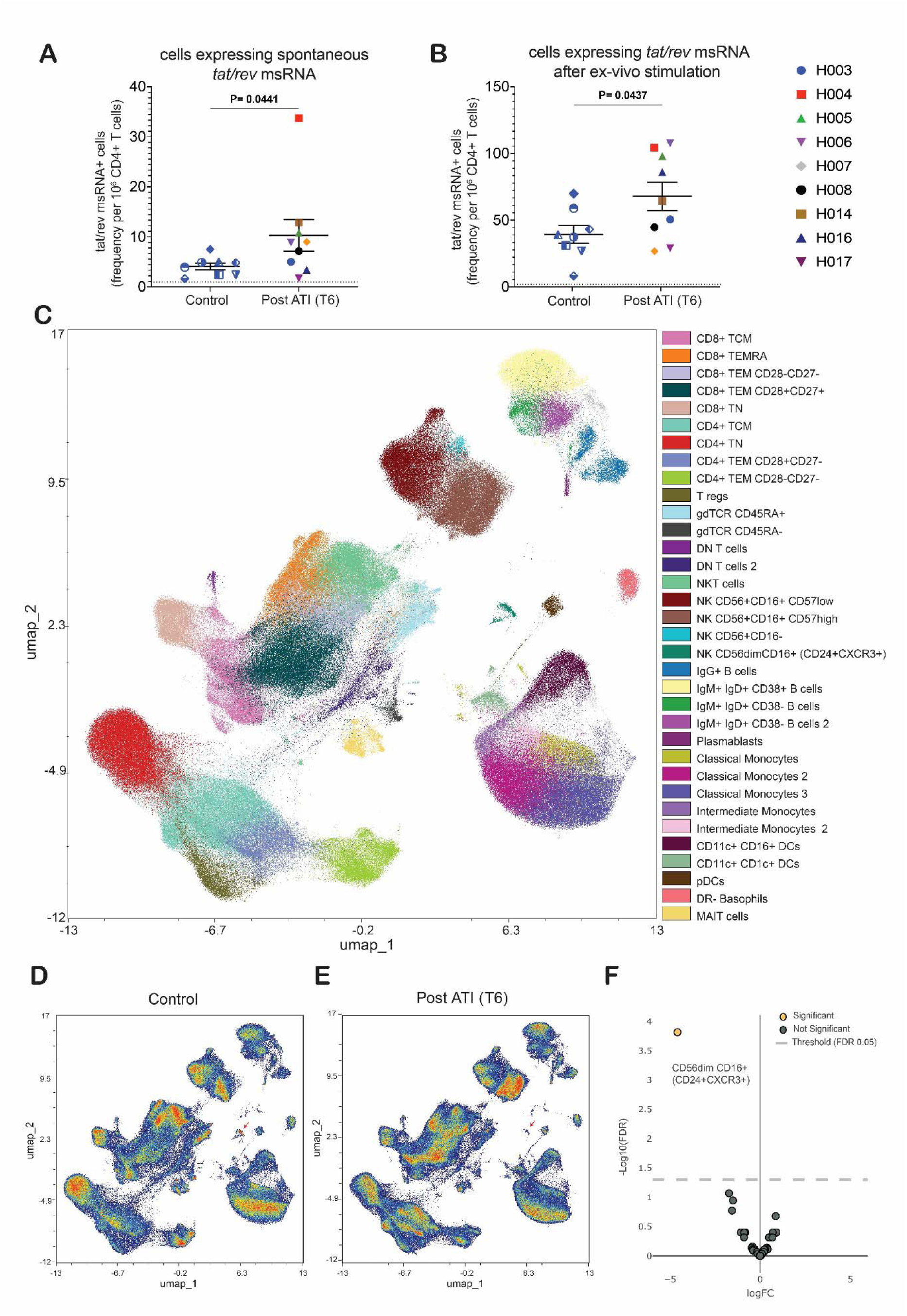
Inducible HIV reservoir and high dimensional flow cytometry analysis of PBMCs from historical controls and post-ATI participants. **A)** Comparison of inducible HIV reservoir using samples obtained from historical HIV+ controls (n=8) and post-ATI participants (n=9) at timepoint 6. CD4+ T cells spontaneously expressing tat/rev msRNA (in vivo response) and **B)** inducibly expressing tat/rev msRNA after ex vivo treatment with PMA/ionomycin for 12, measured using SQuHIVLa. Unpaired t-test was used to compare the means. Two-tailed p-values are shown. **C)** Cell population SPADE clustering of high dimensional 45-color flow cytometry data visualized with uniform manifold approximation and projection (UMAP) dimensionality reduction. Clusters are annotated based on surface marker expression. Plot shows concatenated files from all samples analyzed (n=17). **D-E)** UMAP plots of data analyzed from historical controls (D) and post-ATI participants (E). Red arrow points to a distinct NK cluster CD56dim CD16+ (CD24+CXCR3+) as shown in C. **F)** Volcano plot showing comparison of cluster abundance in historical controls compared to Post-ATI participants as analyzed by edgeR. Negative fold change shows decrease in cluster abundance in Post-ATI participants compared to historical controls. Analysis was conducted with a FDR of 0.05.

### In-depth profiling of the immune compartment a decade following ATI reveals discrete signatures in innate and adaptive subsets

While we observed stable viral re-suppression and immune reconstitution (CD4/CD8 ratios) more than a decade after resuming ART (Fig. 1 B-D), the long-term impact of vaccination and subsequent extended viral rebound on immune architecture and inflammation remains incompletely understood, especially in context of the observed increase in the inducibility of tat/rev msRNA expressing cells. To define whether prolonged ATI leaves an immunological imprint, we performed high-dimensional 45-color spectral flow cytometry on peripheral blood mononuclear cells (PBMCs) from post-ATI participants at T6 and matched historical HIV+ controls (Supplementary Figure 2). Using SPADE clustering algorithm and UMAP-based dimensionality reduction, we identified discrete immune cell clusters (Supplementary Table 2, Supplementary Figure 3A) spanning major lymphoid and myeloid compartments (Fig. 3C). UMAP projections revealed no major phenotypic separation between the two groups, except within natural killer (NK) cell subsets (Fig. 3C-E). Volcano plot analysis of cluster abundances using edgeR (FDR < 0.05) revealed a marked reduction in the frequency of a distinct population of NK cells in the post-ATI group (Fig. 3F, Supplementary Figure 3A-E). This population corresponds to a cluster enriched for CD56dim CD16+ NK cells, characterized by differential expression of CD24+ and CXCR3+ relative to other NK cell clusters (Supplementary Figure 3A-E, Supplementary Table 2).

We next investigated whether functional phenotypes of memory T cells and NK after ATI are different from historical controls by profiling expression of key exhaustion and activation markers across memory T cell and NK populations in post-ATI (T6) participants and historical HIV+ controls. Supervised multivariate analysis using Significance Analysis of Microarrays (SAM; FDR < 0.01) revealed significant differences in the expression pattern of several markers across memory CD8⁺ and CD4+ T cell subsets (Fig. 4A-B). In memory CD8+ T cells, we observed significant upregulation of markers CXCR3 and CD25 across multiple CD8+ T cell memory compartments of post-ATI participants, including TEM CD28+CD27+ and TEMRA cells (Fig. 4C-D, q-value < 0.05). This pattern is also observed In CD4+ memory T cells of post-ATI participants, with similar upregulation of homing and activation markers CXCR3, CCR5, CCR6, KLRG1, and CD25 in effector memory subsets (Fig. 4B, 4E-F). In addition, memory CD8+ T cell subsets show a reduced surface expression of C-type lectin receptor CD161 and differentiation marker CD57 (Fig. 4F), a phenotype previously associated with untreated HIV infection (42, 43). Interestingly, memory CD8+ and CD4+ T cell subsets in post-ATI participants have a modest but significant (q-value < 0.05) decrease of exhaustion/chronic activation-associated markers CD38 and TIGIT (Fig. 4F).

**Fig. 4.**
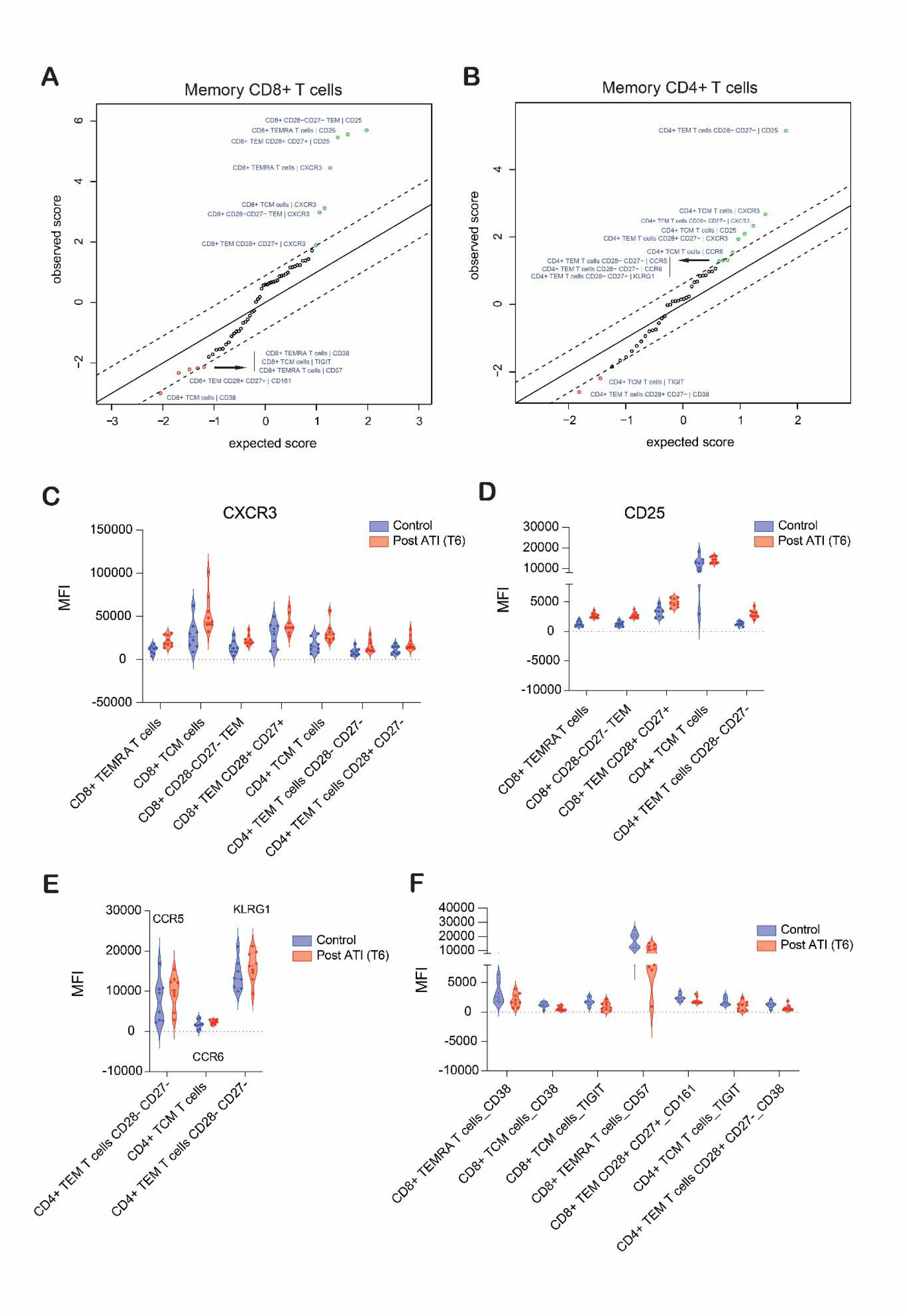
Multivariate differential analysis of T cell exhaustion and activation marker expression in memory T cell clusters. **A-B)** SAM plots depicting differences in median fluorescence intensity of exhaustion and activation markers (CD25, CD38, CD39, HLA-DR, TIGIT, TIM-3, KLRG1, CCR5, CCR6, PD-1, CD57, CXCR3, CXCR5, CD161) in memory CD8+ (A) and CD4+ (B) T cell clusters. The diagonal line represents alignment of observed and expected scores. Colored points outside of the diagonal line represent a significant increase (green) or decrease (red) in median fluorescence intensity of exhaustion and activation markers of Post-ATI participants compared to historical HIV+ controls. Analysis was conducted with 500 permutations and a FDR of 0.01 to ensure robustness. **C-F)**. Violin plots showing mean fluorescence intensity (MFI) values of up- or downregulated activation and exhaustion markers across memory T cell populations identified in the SAM analysis. Plots are grouped based on significant upregulated markers, including CXCR3 (C), CD25 (D), CCR5, CCR6 and KLRG1 (E), as well as downregulated markers (F).

When analyzing NK cell clusters, SAM plots showed no major shift in activation or exhaustion profiles of these subsets, except a moderate downregulation (q-value = 0.22) of markers CD337, CD161, TIGIT, and TIM-3, mostly on CD24+CXCR3+ NK subset in the post-ATI group compared to controls (Supplementary Fig. 4A).

Lastly, we set out to investigate whether the phenotypic differences observed in memory T cell subsets correlate with HIV-1 reservoir dynamics, prompted by a previous study pointing towards CXCR3+ memory CD4+ T cell subsets as a unique cellular reservoir enriched for inducible replication-competent virus (44). Consistent with this data, we observed a significant correlation between CXCR3+ expression on the surface of central memory CD4+ T cells and inducible tat/rev msRNA+ cells (Fig. 5A). This data suggests that CXCR3+ expression may serve as a phenotypic biomarker of the inducible HIV-1 reservoir with potential implications for ATI and viral rebound monitoring.

**Fig. 5.**
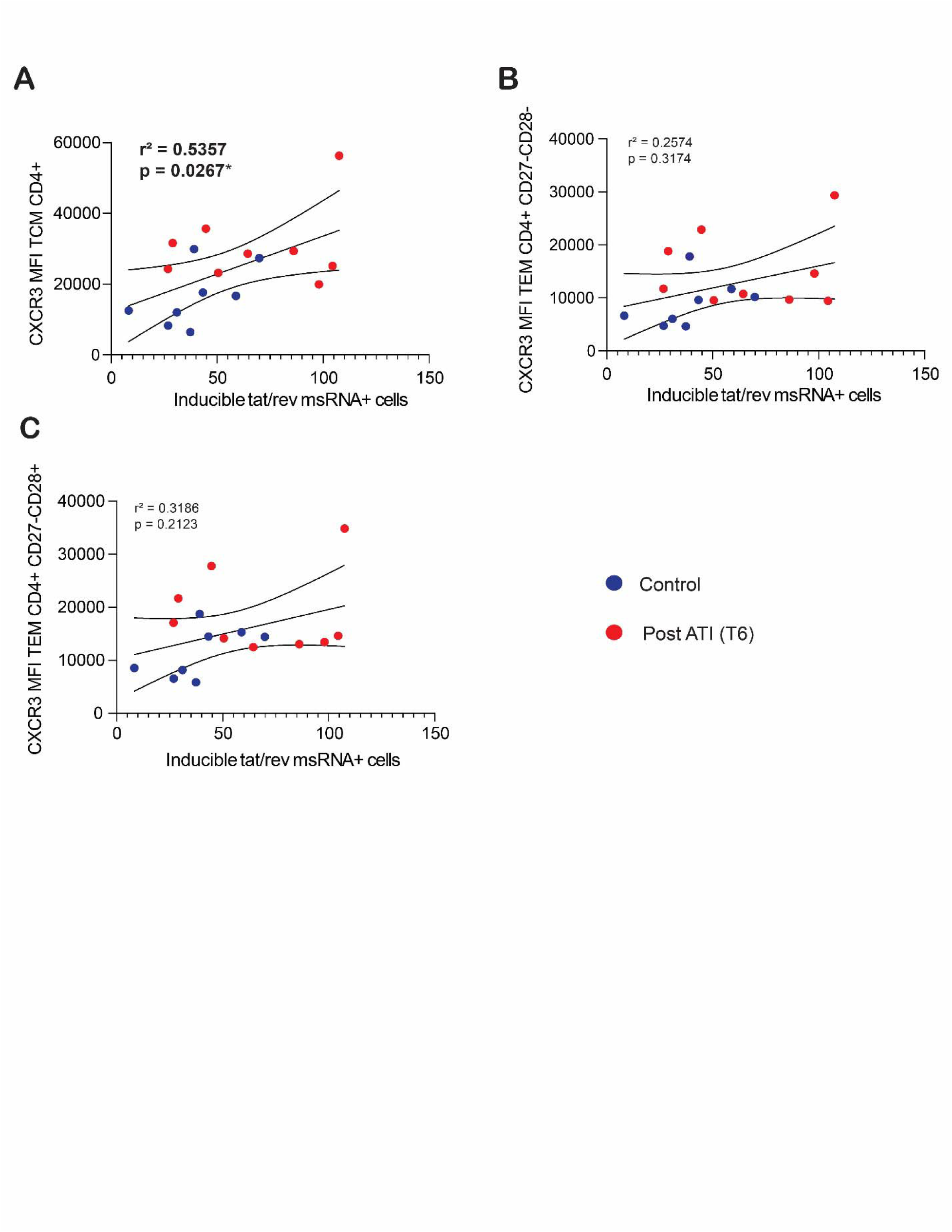
Relationship between T cell subsets expressing CXCR3 and inducible *tat/rev* msRNA+ cells. **A-C)** Linear regression model for CXCR3 mean fluorescent intensity in TCM CD4+ (A), TEM CD4+ CD27+CD28-(B) and TEM CD4+ CD27-CD28-(C) clusters and inducible tat/rev msRNA+ cells measured by SQuHIVLa. Coefficient of correlation and statistical significance was calculated for A (data normally distributed) and Spearman correlation for B and C (data not normally distributed).

## DISCUSSION

In this study, we present a uniquely long-term immunovirological follow-up of individuals who underwent prolonged analytical treatment interruption (ATI) during a dendritic cell (DC)-based HIV-1 vaccine trial, offering a rare opportunity to assess the durable consequences of an ATI more than a decade after ART resumption. This work integrates high-dimensional phenotyping, deep HIV-1 reservoir characterization, and historical comparator analysis to understand the impact of ATI on the latent HIV-1 reservoir and immune architecture.

While ART has proven effective at suppressing viral replication, the reactivation of latent HIV-1 reservoirs following treatment interruption remains a significant barrier to cure. ATI protocols, though essential for assessing cure strategies, carry the risk of viral rebound and potential expansion of the viral reservoir. Our data demonstrate that this expansion may not be limited to the acute viral rebound phase. Indeed, we observed a significant increase in the size of the inducible latent HIV-1 reservoir—defined by the capacity of CD4+ T cells to express tat/rev multiply spliced RNA upon ex vivo stimulation, more than ten years after ATI. This expansion in the inducible latent HIV-1 reservoir, despite sustained virologic suppression and therapeutic ART levels, was confirmed through both longitudinal, intra-individual analysis, and cross-sectional comparison to a matched control group of men with HIV-1. Importantly, the expansion was restricted to the inducible HIV-1 reservoir; post-ATI levels of total and intact proviral DNA measured by IPDA were overall comparable to baseline (Pre-ATI), although 4/9 participants (44.4%) showed a notable upward trend in intact proviral DNA. On the other hand, transcription-competent reservoir cells (gag-pol vRNA+) were remarkably stable. These findings reinforce emerging evidence that HIV-1 reservoir quality, specifically the inducibility and transcriptional activity of proviruses, may be more relevant than total proviral DNA quantity in predicting viral rebound risk and informing HIV-1 cure strategies. Indeed, earlier studies by Fischer et al. demonstrated tat/rev msRNA expression concomitantly occurring with the re-emergence of PBMC-associated HIV-1 particles (i.e., cellular viral rebound) after two weeks of structured ART interruption, which was predicted by during-therapy levels of nef msRNA transcripts in PBMCs (45). More recently, De Scheerder et al., showed an increase in tat/rev RNA transcripts, amongst elevated expression of restriction factors (SLFN11 and APOBEC3G), after ATI prior to viral rebound, indicating the use of these restriction factors and tat/rev RNA as potential biomarkers for imminent viral rebound (25).

Beyond the viral reservoir, our results indicate that prolonged ATI durably imprints on immunophenotypic profiles. Using 45-color spectral flow cytometry and unsupervised clustering, we longitudinally defined ATI-associated immune signatures compared to historical controls. Overall, UMAP projections revealed no major differences in distribution or clustering of immune subsets between groups, consistent with immune reconstitution (Figure 1). However, we did observe a significantly lower frequency of a CD56dim CD16+ NK cluster in T6 samples from post-ATI participants compared to control samples. This cluster was enriched for CD56dim CD16+ NK cells that differentially express high levels of markers CD24+ and CXCR3+ compared to other NK cell clusters. The elevated expression of CXCR3+ suggests a migratory phenotype within this population (46). Interestingly, CD24 is a characteristic marker of B cells and is not typically expressed on NK cells, making this cluster phenotypically distinct. As the functional characteristics and biological relevance of this population are unclear, further studies are warranted to clarify its role in the post-ATI immune landscape. In a previous study investigating the impact of DC vaccination on NK cells in the DCTRN trial participants (47), Laeremans et al., did not observe significant changes in the frequencies of total NK cells over the different time points of the trial (PreVac: prior to vaccination (baseline); Vac2R+Rw1: 1Rweek after the second vaccination; ATIR+R4w and ATIR+R16w: 4 or 16Rweeks after ATI). However, a significant decrease in the CD56dimCD16−subset together with an increase in the CD56dimCD16+ subset was observed at Vac2R+R1w compared to PreVac. The frequencies of the CD56dimCD16− and CD56dimCD16+ subsets did not return to PreVac levels after ATI, suggesting a sustained impact of vaccination and ATI on the distribution of these NK cell subsets. Due to limited quantities or unavailable PBMCs at baseline (T0, PreVac), we were unable to determine if the altered frequency of CD56dimCD16− and CD56dimCD16+ NK cells persisted at T6 (after ART resumption and long-term viral re-suppression). We were also unable to bridge additional insights from the two studies due to non-overlapping time points (longitudinal vs cross-sectional) and differences in the markers analysed as well as the control samples used i.e., HIV-1 negative (Laereman’s study) vs chronic HIV-1 after long term, uninterrupted ART (this study).

In CD4+ and CD8+ memory T cells, we observed a complex phenotypic profile: increased surface expression of homing and activation markers (CCR5, CCR6, CXCR3, CD25, KLRG1), decreased expression of mucosal marker CD161 and differentiation marker CD57, and a partial reduction in classical exhaustion and chronic activation markers (TIGIT, CD38). Similar shifts in CXCR3, CD161 and CD57 expression have been reported in progressive HIV infection and in cancer, where they are associated with altered effector function and immune dysregulation (42, 48, 49). A decreased expression of TIGIT and CD38 indicates that memory T cell compartments of post-ATI participants, rather than an overall exhaustion profile, present a migratory/activated profile. However, further assays are needed to determine if these phenotypic changes present in the post-ATI participants associate with a functional phenotype.

Notably, out of all the markers differentially expressed, activation and tissue homing receptor CXCR3+ was observed to be highly and significantly upregulated in both memory CD4+ and CD8+ T cell subsets of post-ATI participants. CXCR3 is a chemokine receptor expressed on T cells upon activation and plays a crucial role in trafficking and effector responses of memory T cells (50). In people with HIV, CXCR3+ expression is increased in populations of circulating memory CD4+ and CD8+ T cells and gut mucosal CD4+ T cells compared to healthy individuals (51, 52). An increase of CXCR3 expression on subsets of circulating memory T cells post-ATI may indicate that more T cells in these people with HIV are migrating into tissues and this could be due to ongoing tissue inflammation and active immunosurveillance.

Interestingly, a recent study showed that circulating CXCR3+ memory CD4+ T cells are enriched for inducible replication-competent provirus and contribute to the latent HIV-1 reservoir pool (44). A deeper look into our data revealed that, in our cohort, CXCR3+ expression on the surface of central memory CD4+ T cells positively correlates with inducible tat/rev msRNA expression. Importantly, this significant correlation is not observed in effector memory subsets, indicating a selective association with central memory CD4+ T cells, also considered the main cellular reservoir of HIV-1 proviruses (53). These findings suggest that CXCR3+ expression on memory CD4+ T cells could serve as a functional biomarker for inducible HIV-1 provirus, and monitoring CXCR3+ memory subsets during ATI could offer valuable insights into HIV-1 reservoir dynamics and viral rebound. Of note, we previously found that an increase in CXCR3 ligands coincides with HIV reactivation during ATI in this study population (54).

Our study offers a practical framework for integrating high-dimensional immunovirological profiling into HIV cure trial design. The combination of inducible HIV reservoir assays (such as SQuHIVLa) and multivariate immunophenotyping can provide a richer picture of post-ATI immune health than conventional plasma viral load or CD4⁺ T cell counts alone. Nevertheless, several limitations merit discussion. The sample size is small, owing to the rarity of long-term ART-treated ATI participants with banked longitudinal samples, and may underpower some subgroup analyses. The use of historical comparators, while well-matched, lacks concomitant control of unmeasured variables such as HIV-1 total and intact proviral DNA and gag-pol RNA+ cells. Furthermore, in the absence of viable clinical samples at key phases (pre-ATI, during ATI and upon resuming ART), critical analyses to measure immunological shifts in relation to the vaccine or early during ATI, prior to viral rebound, are not possible with the current analytical tools. However, a previous study noted profound and durable changes in the blood transcriptome in these individuals after vaccination and subsequent ATI (33).

From a translational perspective, our data argue for a cautious, individualized approach to ATI in cure trials, with extended monitoring periods and integration of functional immune metrics. While short-term ATI protocols are increasingly standardized, the long-term consequences remain under-characterized. In our study, the persistence of inducible HIV-1 reservoir expansion and memory immune cell alterations, even in the context of successful viral re-suppression, suggests that the DC vaccination and (prolonged) ATI may have had immunologic costs that are not immediately apparent.

Ultimately, our study does not argue against the use of ATI in HIV-1 cure research; rather, it highlights the need to embed longitudinal immunologic analyses into trial designs. Future efforts should explore strategies to minimize HIV-1 reservoir expansion and preserve immune function during ATI phases, possibly through adjunctive therapies. Our findings also support the development of phenotypic biomarkers (e.g., TIGIT+/CXCR3+/CD38+ memory T cells) that could inform participant selection or stratification in future interventional trials and guide ATI risk.

In conclusion, this study provides a comprehensive, long-term view of the immunologic and virologic consequences of (prolonged) ATI, revealing persistent remodeling of the inducible HIV-1 reservoir and immune architecture. By linking molecular HIV persistence markers with phenotypic immune shifts, we underscore the importance of integrating long-term safety monitoring and immune profiling into the roadmap toward HIV remission.

## MATERIALS AND METHODS

### Ethics statement

Participants of the DCTRN trial provided written informed consent prior to inclusion in accordance with the regulations of the institutional Medical Ethics Committee at Erasmus Medical Center, Rotterdam (MEC-2005-227 and MEC-2012-583). The remaining material from the DCTRN trial was stored in a registered biobank, which was approved by the Erasmus Medical Center Medical Ethics Committee (MEC-2022-0060). The HIV biobank is described at https://eheg.nl/en/Our-Projects/HIV-Biobank. Samples used as controls were obtained from participants of studies at Erasmus Medical Center: ExVivo study, HIV Biomarker study and LUNA clinical trial, with approvals from the institutional Medical Ethics Committee at Erasmus Medical Center (MEC-2012-583, MEC-2016-148, and MEC-2017-476, respectively).

### Plasma HIV-1 RNA monitoring

Quantification of plasma HIV-1 RNA was performed on three different platforms available at various longitudinal timepoints during DC-immunotherapy, at treatment interruption and on ART. Between 2002 and 2008, quantification was performed using the Roche Amplicor HIV-1 RNA monitor test (version 1.5) (LOQ 50 copies/mL). After 2009 plasma HIV-1 RNA was quantified using the COBAS TaqMan Assay (Roche); LOQ 20 copies/mL. As of 2021, plasma HIV-1 RNA was quantified using the Aptima HIV-1 Quant Dx assay; LOQ 30 copies/mL.

### Clinical sample collection and cell isolations

Cryopreserved PBMCs obtained from large blood draws or leukapheresis material collected from the study participants during the trial phase and post intervention were used for this study. Upon thawing PBMCs, CD4+ T cells were isolated by negative magnetic selection using the EasySep Human CD4+ T cell Enrichment kit (StemCell Technologies) according to the manufacturer’s instructions. The isolated CD4+ T cells were either directly used to quantitate HIV-1 DNA/RNA levels or treated with 100 ng/mL phorbol 12-myristate 13-acetate (PMA) and 1 μg/mL ionomycin to enhance viral reactivation.

### Intact Proviral DNA Assay (IPDA)

The levels of intact proviral DNA were assessed using lysed extracts of CD4+ T cells. Nucleic acid was isolated from at least 2 x 10^6^ CD4+ T cells using the PCI extraction method. Duplex digital PCR (dPCR) was performed on an Absolute Q digital PCR platform (Thermo Fisher Scientific) using primer and probe sets targeting the packaging signal (Ψ) and Envelope gene (Env), following the manufacturer’s instructions and adapted intact proviral DNA assay protocol(7, 55, 56). Duplex HIV-1 dPCR reaction mix consisted of 1.8 μL Absolute Q DNA Digital PCR Master Mix 5X (Thermo Fisher Scientific), 1 μL packaging signal (Ψ) target FAM-probe (10x), 1 μL Env target HEX-probe (10x), 700 ng gDNA, diluted in nuclease-free H2O to a final reaction volume of 9 μL. The dark competition probe for APOBEC3G hypermutations in the ENV region was omitted this reaction setup. Measurements of the cellular gene ribonuclease P/MRP subunit p30 (RPP30) in a replicate well were used to calculate the frequency of CD4+ T cells. The duplex RPP30 dPCR was set up similarly using RPP30 primer and probe sets and 7 ng gDNA input. The primer and probe sequences used are provided below.

IPDA primers and probes:

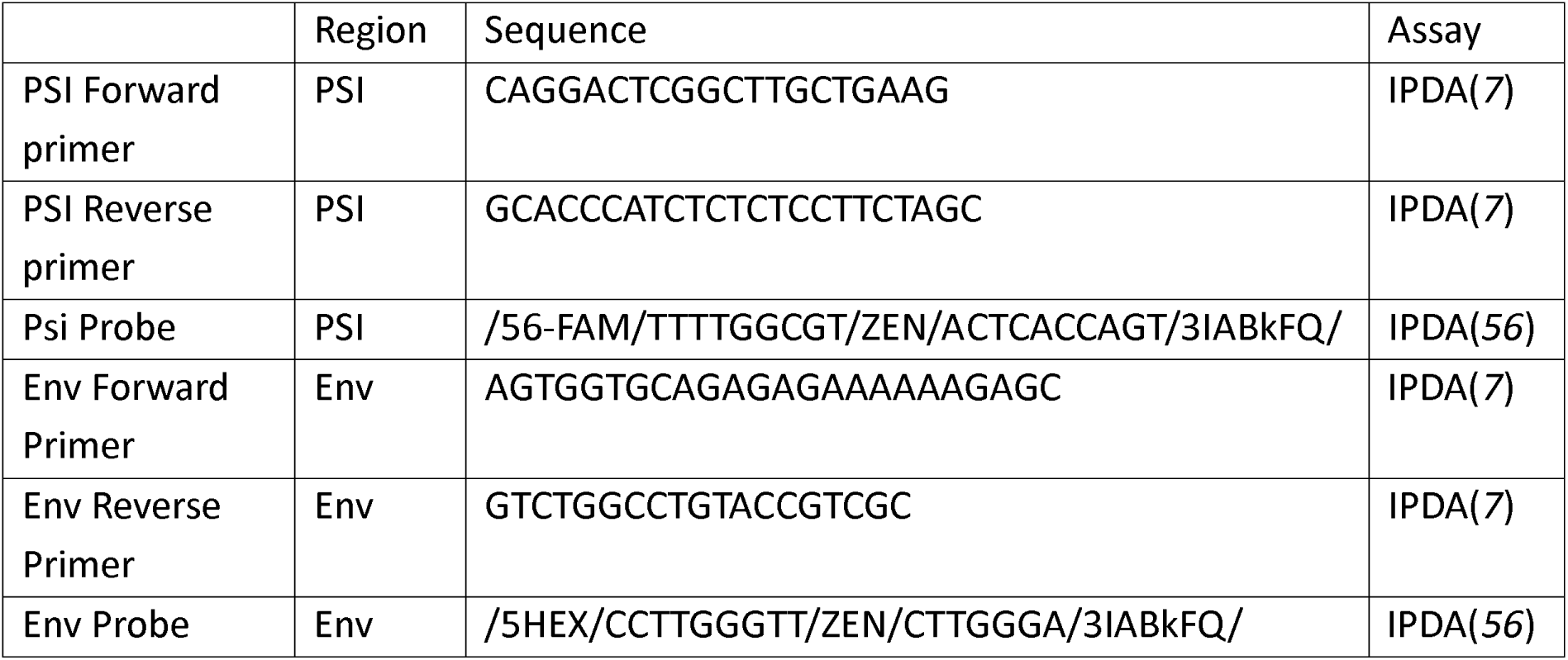

### FISH-flow

The frequency of cells expressing gag-pol viral RNA (vRNA) and p24 protein were assessed using the Primeflow RNA Assay (Thermo Fisher Scientific) following the manufacturer’s instructions and as described previously (37, 57). Briefly, CD4+ T cells were isolated from people with HIV donor PBMCs by negative magnetic selection using the EasySep Human CD4+ T cell Enrichment kit (StemCell Technologies) according to the manufacturer’s instructions. A minimum of 20 x 10^6^ CD4+ T, resuspended at 1.5 x 10^6^ cells/mL in RPMI-1640 medium supplemented with 10% FBS, 100 μg/mL penicillin-streptomycin and Raltegravir (3 μM) and rested for at least 4 hours in a humidified incubator at 37°C and 5 % CO_2_. Cells were then treated with 100 ng/mL PMA and 1 μg/mL ionomycin to enhance viral reactivation. Eighteen hours post stimulation, cells were washed, pellets resuspended in PBS, and counted using an automated cell counter (Countess II, Thermo Fisher Scientific). At least 10 x 10^6^ cells, with >55% viability, were subjected to the FISH-flow assay. CD4+ T cells were first stained in Fixable Viability dye 780 (Thermo Fisher Scientific) for 20 min at room temperature (1:1000 dPBS) and then fixed and permeabilized using reagents from the Primeflow RNA assay kit according to manufacturer’s instructions. After permeabilization, the cells were stained with two p24 antibodies; anti-p24 KC57-FITC (Beckman Coulter, 6604667) and anti-p24 28B7-APC (MediMabs, MM-0289-APC), incubated in the dark for 30 min at room temperature, and an additional 30 minutes at 4°C. Cells were then washed and resuspended in RNA wash buffer with RNAsin. mRNA was labelled with a set of 40 probe pairs against the GagPol region of vRNA (catalogue number: GagPol HIV-1VF10-10884, Thermo Fisher Scientific) diluted 1:5 in the probe diluent provided in the kit. Target mRNA hybridization was carried out for 2 hours at 40°C. Samples were washed to remove excess probes and stored overnight, at 4°C, in the presence of RNAsin. Signal amplification was then performed by sequential 1.5-hour, 40°C incubations with the pre-amplification and amplification mix. Amplified mRNA was labelled with fluorescently tagged probes for 1 hour at 40°C. Samples were washed, resuspended in 200-300 μL storage buffer and up to 5×10^6^ cells were acquired on a BD LSR Fortessa Analyser. Gates were set using stimulated, uninfected CD4+ T cells. Data were analyzed using the FlowJo V10 Software (Treestar).

### Specific Quantitation of Inducible HIV-1 reservoir by LAMP (SQuHIVLa)

The frequency of cells spontaneously and inducibly expressing tat/rev multiply (MS) spliced HIV-1 RNA was assessed using a SQuHIVLa assay according to the protocol described elsewhere (41). Briefly, 2-5 x 10^6^ total CD4+T cells were resuspended (1.5 x 10^6^ cells/mL) in culture RPMI-1640 medium supplemented with 10% FBS and 100 μg/mL penicillin-streptomycin, and rested for 6 hours in a humidified incubator at 37 °C and 5 % CO_2_. After incubation, some cells were left untreated and directly used to determine basal viral RNA expression or treated with 100 ng/mL PMA and 1 μg/mL ionomycin for 12 hours to induce viral reactivation. Cells from either condition were washed twice in RPMI 1640 media with 3% FBS, then resuspended in PBS and counted using an automated cell counter (Countess II, Thermo Fisher Scientific). A minimum of 0.5 x 10^6^ CD4+ T cells, with > 50 % viability, after PMA/ionomycin treatment were analyzed per SQuHIVLa assay performed on a CFX96 Touch Real-Time PCR Detection System thermocycler (BioRad) as described elsewhere (41).

### One-step Tat/rev Induced Limiting Dilution Assay (TILDA)

The frequency of cells expressing tat/rev multiply spliced (ms) HIV-1 RNA was assessed using a modified TILDA protocol (58) i.e. by direct amplification of tat/rev msRNA using a one-step RT-qPCR approach. Briefly, 2 - 5 x 10^6^ CD4+ T cells isolated from people with HIV donor PBMCs were rested and treated with PMA/ionomycin as described for the other inducible reservoir assays. A minimum of 0.5 x 10^6^ CD4+ T cells, with >50 % viability, after treatment with PMA/ionomycin were analyzed per assay. CD4+ T cells were added in limiting dilution, ranging from 3 x 10^4^ cells - 3 x 10^3^ cells (22-24 replicates), to a white 96-well qPCR plate (Biorad) containing 20 μL One-step RT-qPCR reagent mix; 5 μL Luna Probe One-Step RT-qPCR 4X Mix with UDG (No ROX) (New England BioLabs), 0.1 μL tat 1.4 forward primer and 0.1 μL rev reverse primer (both at 100 μM), 0.8 μL HIV tat/rev probe (5 μM), and 14 μL nuclease free water to a final reaction volume of 25 μL. The primer and probe sequences used, as published(58). The qPCR plates were sealed, briefly centrifuged at 600 x g for 1 min, and a CFX96 Touch Real-time PCR instrument (Biorad) was used to run the following thermocycling program; Carry over prevention step at 25°C for 30 seconds, reverse transcription at 55°C for 10 minutes, initial denaturation at 95°C for 3 minutes followed by 50 cycles of denaturation at 95°C for 10 seconds and extension at 60°C for 1 minute. The amplification curves in all positive wells at each dilution manually inspected after each run and the number of positive wells were used to determine the frequency of cells positive for tat/rev msRNA using the maximum likelihood method.

### In-depth immunophenotyping of the major cell subsets present in human peripheral blood

PBMCs from T6 participants and historical HIV+ controls, matched by age, sex and time on ART (Supplementary Fig. 2), were thawn and stained with a 45-color flow cytometry panel and data was read on a 5-laser Aurora spectral flow cytometer (Cytek Biosciences, CA). Staining of the samples was performed following the OMIP-109 platform with minor modifications (59). Specifically, we replaced the live/dead stain for anti-Annexin V conjugated antibody (catalogue number: A23202, Thermo Fisher Scientific) to exclude both dead and apoptotic cell populations and added 2.5 mM CaCl_2_ to all buffers. In addition, to better define a CD20+CD19+ population, we added anti-CD20 antibody as a last step of the sequential staining for 5 minutes at room temperature, before staining with the full remaining antibody cocktail. Spectral unmixing and compensation check was performed in SpectroFlo (version 2.2.0). Flow cytometry data clean-up was performed using the flowAI algorithm embedded in the OMIQ data analysis software (www.omiq.ai). Our downstream data processing workflow included scaling adjustments, sub-sampling (max equal) and manual gating to define live populations of single cells. For high-dimensional analysis, we used SPADE (60) algorithm for clustering and UMAP (61) projections for dimensionality reduction and 2D visualization of defined clusters. For analysis of cluster abundance differences between groups, we used edgeR algorithm (FDR <0.05) and volcano plots for visualization. For multivariate statistical analysis of the median fluorescent expression of activation and exhaustion markers in specific clusters, we used Significant Analysis of Microarrays (SAM) plot algorithm with FDR <0.01. All programs used for analysis are embedded within the OMIQ data analysis software.

### Statistics and reproducibility

Data are presented as median with Inter Quartile Range. Spearman rank correlation coefficients were calculated for non-normally distributed variables. Two-tailed P values were calculated. P < 0.05 was considered statistically significant. For the inducible viral reservoir quantification data below the limit of detection, the arbitrary value of 0.5 cells/million were used to plot the data but were not included in statistical analyses. Likely outlier data points were identified using ROUT(Q=10%) and excluded from statistical analyses. Mixed effects model analyses and Tukey’s multiple comparison test were used to compare viral reservoir frequencies across timepoints. Adjusted P values were calculated. P < 0.05 was considered statistically significant. Biological replicates (that is, samples at longitudinal timepoints) were measured in most experiments except for inducible HIV reservoir quantification given the limited availability or quality of clinical material (particularly baseline samples). Technical replicates (that is, repeated measurements of the same sample) were as follows: for intact proviral DNA quantification by dPCR, at least two technical replicates; for FISH-flow, experiments were not technically replicated given the limited clinical material; for SQuHIVLa and 1-step TILDA, at least two independent experiments were performed where clinical material was sufficient. Prism 9 for Mac (GraphPad Software) was used to generate graphs and to perform all statistical tests.

## LIST OF SUPPLEMENTARY MATERIALS

Supplementary Fig. 1 to 4

Supplementary Table 1 and 2

## Data Availability

All data needed to evaluate the results and conclusions in this article are provided in the main text, Methods section and Supplementary data section. Additional data related to the study may be requested from the lead contact T. Mahmoudi. Unique biological material, where available, can be obtained upon request.

## ACKNOWLEDGEMENTS

We express our deepest gratitude to all the study participants for their valuable contribution to the DCTRN trial in 2006-2009 and to the current study, without whom this research would not have been possible. We also thank the internist-infectious disease specialists at Erasmus MC for their assistance in recruiting the study participants. Leukapheresis procedures and sample logistics were greatly supported by Erasmus MC HIV research nurses and clinical laboratory staff. C.L. received funding from the Dutch Aidsfonds (grants P-60602 and P-263); SR received funding from Dutch Aidsfonds (grant P-53302); C.R. received funding from Dutch Aidsfonds (grant P-53601) and MRACE (EMC2020-083); R.A.G. received funding from Dutch Aidsfonds (grant P-60804) and Horizon Europe (grant 681032); TMa received funding from Health Holland (grants LSHM19100-SGF and EMCLSH19023) and ZonMW (grant 40-44600-98-333) and a Building Synergistic Infrastructure for NL4Cure grant.

## AUTHOR CONTRIBUTIONS

Conceptualization, C.L, and T.Ma; Methodology, C.L., T.H., RC., Y.M.M, and T.Ma; Investigation, C.L., T.H., R.C., H.A.B.P., K.S.H.; Data Curation and Formal Analysis, C.L., T.H., R.C., and T.Ma; Data Visualization, C.L., T.H., R.C., and T.Ma., Project Administration, C.L., R.C., H.A.B.P., K.S.H. and T.Ma. Supervision, R.A.G., C.R., Y.M.M., and T.Ma; Material and Funding Acquisition, C.L., S.R., C.R., R.A.G., Y.M.M., and T.Ma., Writing – Original draft, C.L. T.H., and R.C.; Writing – Review and Editing, C.L., T.H., R.C., H.A.B.P., K.S.H., C.R., S.R., J.J.A.vK., D.A.M.C.V., T.M., P.D.K., Y.M.M., R.A.G., and T.Ma.

