## Supplemental Data for "In-depth profiling of the immuno-virological landscape after a decade of sustained HIV-1 suppression following ART resumption"

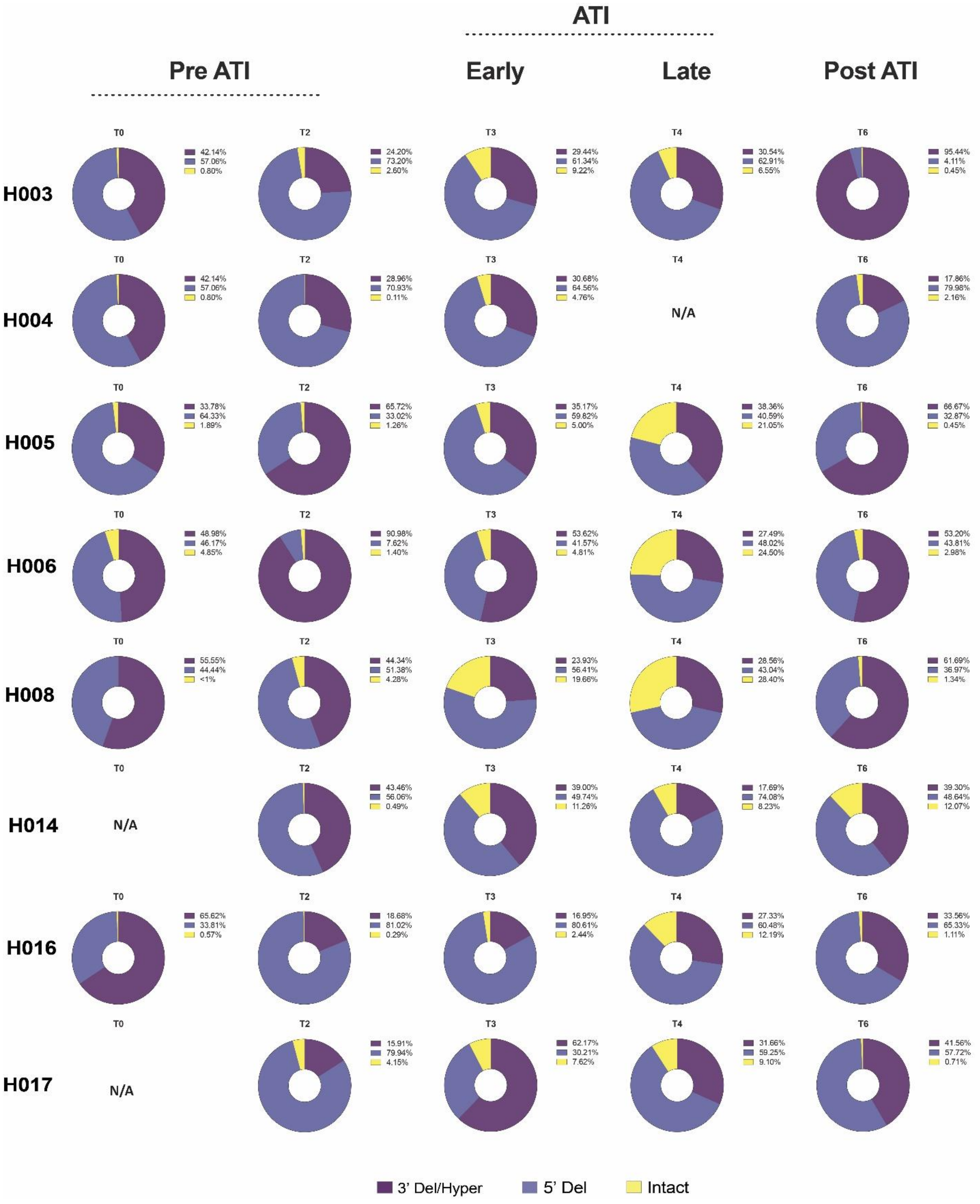

ATI
Pre ATI
Early
Late
Post ATI
H003
T0
42.14%
57.06%
0.80%
T2
24.20%
73.20%
2.60%
T3
29.44%
61.34%
9.22%
T4
30.54%
62.91%
6.55%
T6
95.44%
4.11%
0.45%
H004
T0
42.14%
57.06%
0.80%
T2
28.96%
70.93%
0.11%
T3
30.68%
64.56%
4.76%
T4
N/A
T6
17.86%
79.98%
2.16%
H005
T0
33.78%
64.33%
1.89%
T2
65.72%
33.02%
1.26%
T3
35.17%
59.82%
5.00%
T4
38.36%
40.59%
21.05%
T6
66.67%
32.87%
0.45%
H006
T0
48.98%
46.17%
4.85%
T2
90.98%
7.62%
1.40%
T3
53.62%
41.57%
4.81%
T4
27.49%
48.02%
24.50%
T6
53.20%
43.81%
2.98%
H008
T0
55.55%
44.44%
<1%
T2
44.34%
51.38%
4.28%
T3
23.93%
56.41%
19.66%
T4
28.56%
43.04%
28.40%
T6
61.69%
36.97%
1.34%
H014
T0
N/A
T2
43.46%
56.06%
0.49%
T3
39.00%
49.74%
11.26%
T4
17.69%
74.08%
8.23%
T6
39.30%
48.64%
12.07%
H016
T0
65.62%
33.81%
0.57%
T2
18.68%
81.02%
0.29%
T3
16.95%
80.61%
2.44%
T4
27.33%
60.48%
12.19%
T6
33.56%
65.33%
1.11%
H017
T0
N/A
T2
15.91%
79.94%
4.15%
T3
62.17%
30.21%
7.62%
T4
31.66%
59.25%
9.10%
T6
41.56%
57.72%
0.71%
3’ Del/Hyper
5’ Del
Intact

**Supplementary Fig. 1. Longitudinal HIV-1 DNA dynamics.** Pie charts of intact and defective HIV-1 DNA (5’ Deleted [5’Del] and 3’ Deleted/Hypermutated [3’Del/Hyper]) are shown for each participant. The proportions of intact and defective DNA are shown as percentages of total HIV DNA copies/million CD4+ T cells. Intact HIV DNA was distinguished from defective HIV DNA using the intact proviral DNA assay (IPDA). N/A = not available.

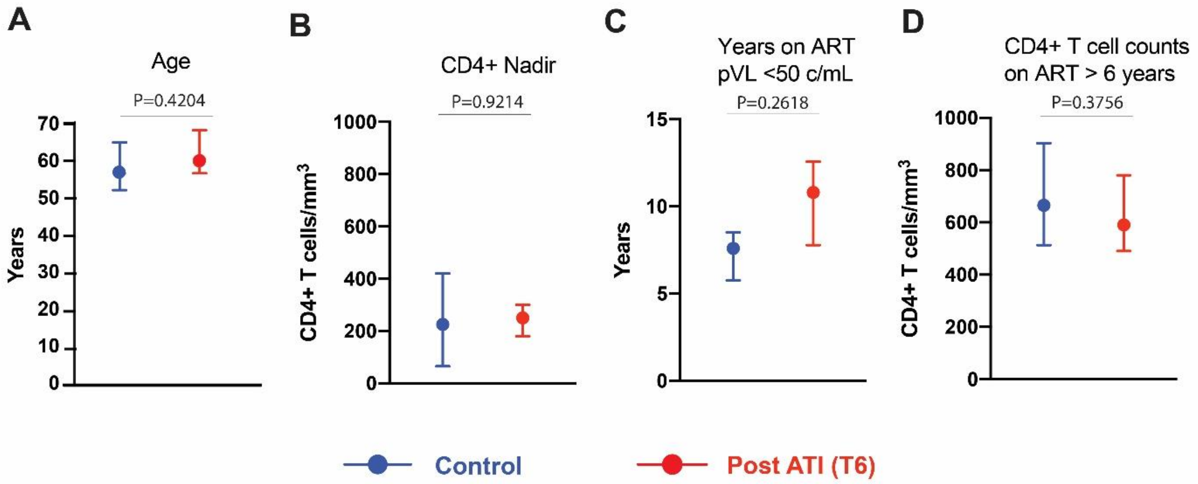

| | Control | Post ATI (T6) |
|---|---|---|
| Sex assigned at birth, n | Male (8/8) | Male (9/9) |
| Age, median (IQR) | 57 (52-65) | 60 (57-68) |
| CD4+ T cell Nadir, median (IQR) | 225 (65-420) | 250 (180-300) |
| Years on ART, median (IQR) | 9.9 (7.1-18.3) | 12.4 (11.9-13.5) |
| Years on ART, <50 c/mL, median (IQR) | 7.6 (5.8-8.5) | 10.8 (7.8-12.6) |
| Last CD4+ T cell count, median (IQR) | 665 (513-903) | 590 (490-780) |

**Supplementary Fig. 2. Clinical characteristics of historical control cohort compared to post-ATI trial participants. A-D)** Comparison by age, CD4 Nadir, years on ART with suppressed plasma HIV RNA below 50 copies/mL and CD4+ T cell counts at time of sampling (>6 years on suppressive ART). The error bars represent median and IQR. The characteristics are summarized in the table. Unpaired t-test was used to compare the means. Two-tailed P values are shown.

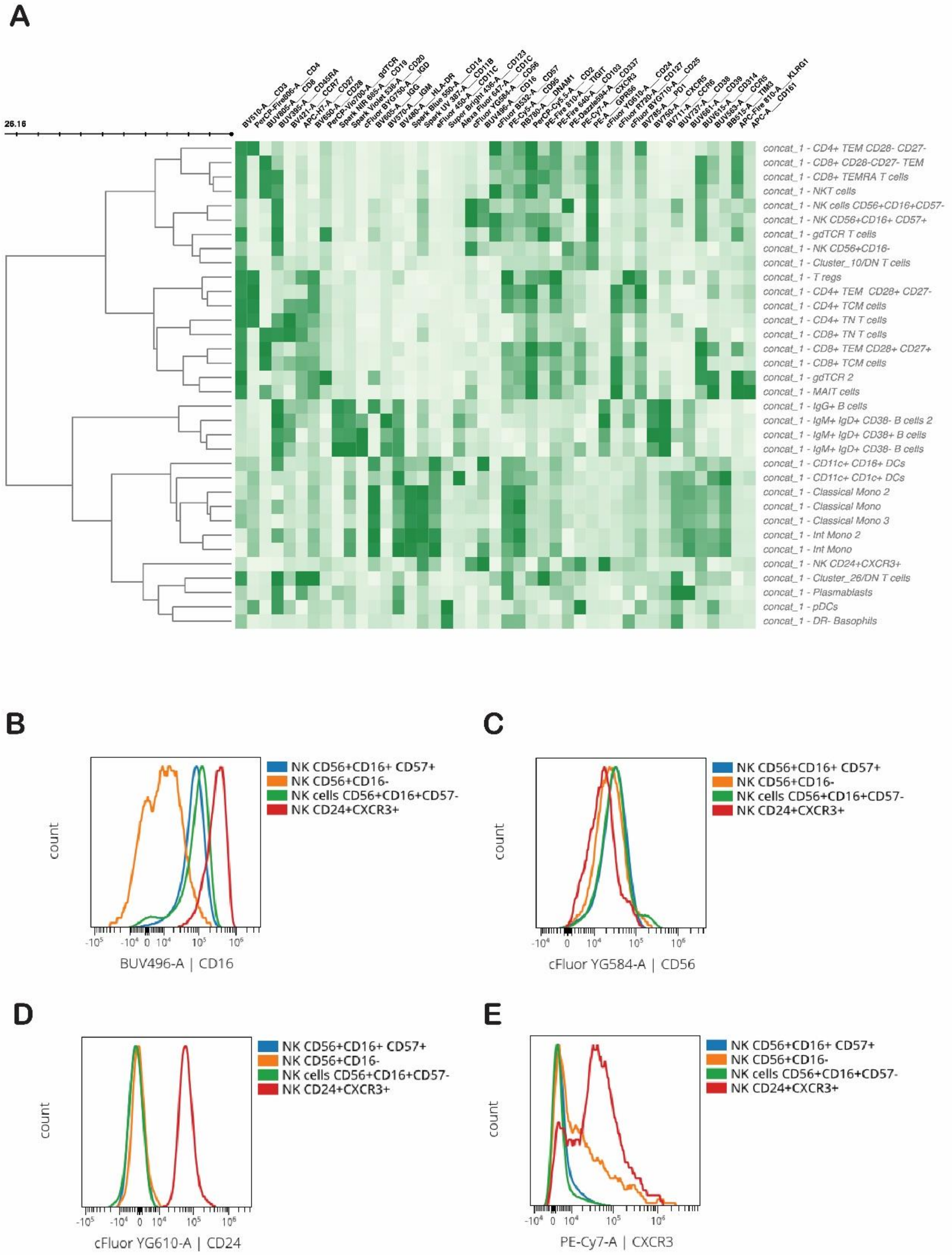
A
26.16
concat_1 - CD4+ TEM CD28- CD27-
concat_1 - CD8+ CD28-CD27- TEM
concat_1 - CD8+ TEMRA T cells
concat_1 - NKT cells
concat_1 - NK cells CD56+CD16+CD57-
concat_1 - NK CD56+CD16+ CD57+
concat_1 - gdTCR T cells
concat_1 - NK CD56+CD16-
concat_1 - Cluster_10/DN T cells
concat_1 - T regs
concat_1 - CD4+ TEM CD28+ CD27-
concat_1 - CD4+ TCM cells
concat_1 - CD4+ TN T cells
concat_1 - CD8+ TN T cells
concat_1 - CD8+ TEM CD28+ CD27+
concat_1 - CD8+ TCM cells
concat_1 - gdTCR 2
concat_1 - MAIT cells
concat_1 - IgG+ B cells
concat_1 - IgM+ IgD+ CD38- B cells 2
concat_1 - IgM+ IgD+ CD38+ B cells
concat_1 - IgM+ IgD+ CD38- B cells
concat_1 - CD11c+ CD16+ DCs
concat_1 - CD11c+ CD1c+ DCs
concat_1 - Classical Mono 2
concat_1 - Classical Mono
concat_1 - Classical Mono 3
concat_1 - Int Mono 2
concat_1 - Int Mono
concat_1 - NK CD24+CXCR3+
concat_1 - Cluster_26/DN T cells
concat_1 - Plasmablasts
concat_1 - pDCs
concat_1 - DR- Basophils
B
NK CD56+CD16+ CD57+
NK CD56+CD16-
NK cells CD56+CD16+CD57-
NK CD24+CXCR3+
count
BUV496-A | CD16
C
NK CD56+CD16+ CD57+
NK CD56+CD16-
NK cells CD56+CD16+CD57-
NK CD24+CXCR3+
count
cFluor YG584-A | CD56
D
NK CD56+CD16+ CD57+
NK CD56+CD16-
NK cells CD56+CD16+CD57-
NK CD24+CXCR3+
count
cFluor YG610-A | CD24
E
NK CD56+CD16+ CD57+
NK CD56+CD16-
NK cells CD56+CD16+CD57-
NK CD24+CXCR3+
count
PE-Cy7-A | CXCR3

**Supplementary Fig. 3**. **Immune Cell Subset Clustering**. **A**) Clustered heatmap of medians for 43 features (excl. CD45 and Annexin V as cells were subsampled on CD45+ Live) for all clusters defined by SPADE (Fig. 3). All 17 samples were concatenated and relative median fluorescence expression was used to annotate the different clusters. **B-E)**. Histogram overlay of CD16 (B), CD56 (C), CD24 (D) and CXCR3 (E) in distinct NK populations in concatenated samples.

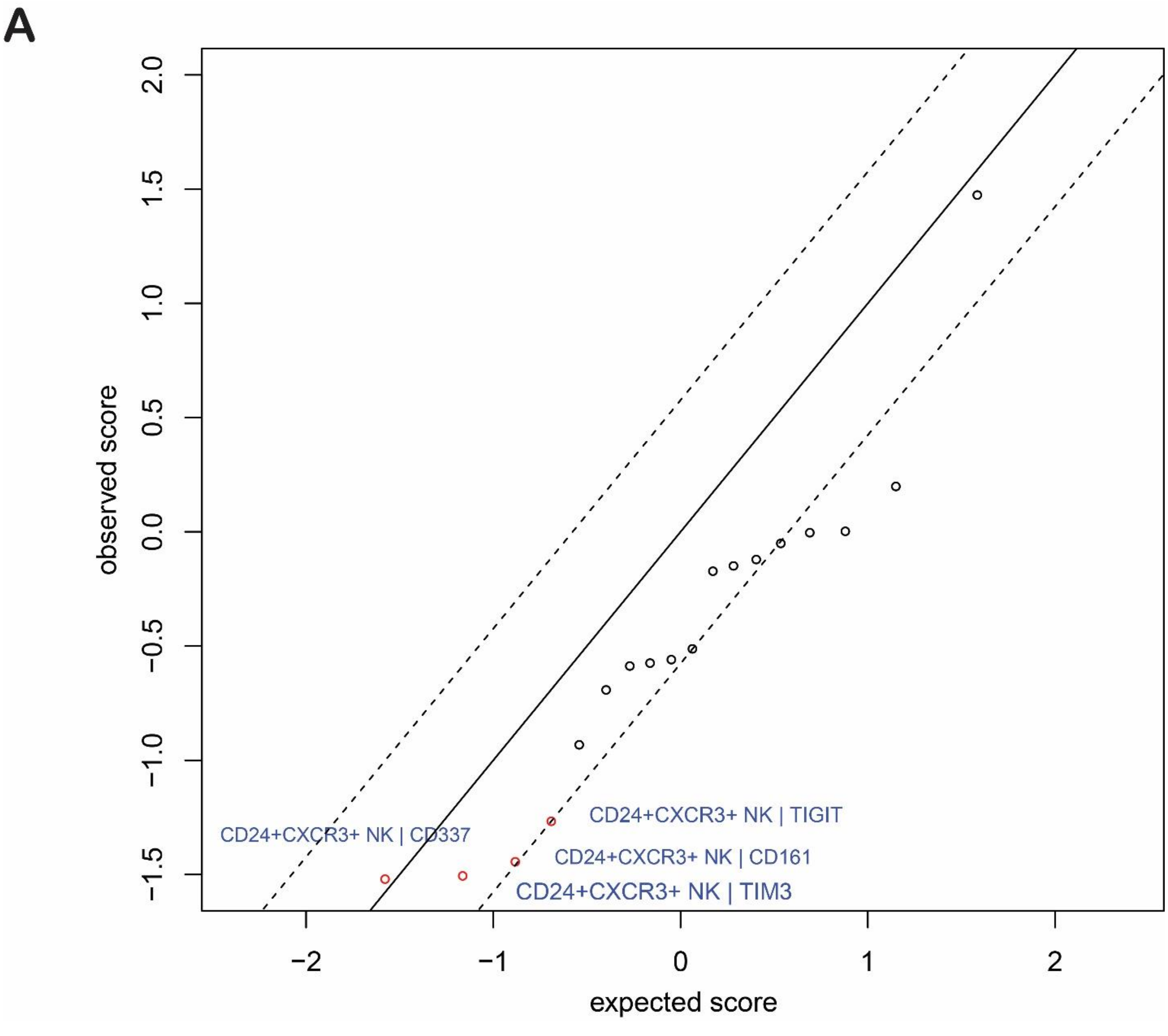

**Supplementary Fig. 4**. **Multivariate differential analysis of exhaustion and activation marker expression in NK cells. A**) SAM plot shows changes in median fluorescence intensity of surface markers PD-1, TIM-3, KLRG1, TIGIT, CD161, CD337 in NK cell clusters. The diagonal line represents alignment of observed and expected scores. Red points outside of the line represent a significant decrease (red) in median fluorescence intensity of said markers.

**Supplementary Table 1** Participant clinical characteristics

| | **Median** | **IQR** | **Range** |
|---|---|---|---|
| Sex assigned at birth | Male (9/9) (100%) | | |
| Age at DC Vaccination (T0) | 45 | 41.5 - 53 | 37 - 56 |
| Age on stable ART post intervention (T6) | 60 | 56.5 - 68 | 52 - 71 |
| Year of HIV Diagnosis | 1997 | 1994 -1999 | 1988 -2000 |
| Time between HIV diagnosis and ART initiation (weeks) | 32 | 18.5 - 198.0 | 2 - 442 |
| Time on ART before vaccination (years) | 9.9 | 7.0 -10.85 | 3.7 - 11.9 |
| Time on ART with undetectable plasma HIV RNA (<50 c/mL) before DC vaccination (years) | 4.8 | 4.6 - 5.0 | 2.8 - 5.3 |
| Peak plasma HIV RNA pre-ART (log10 c/mL) | 5.01 | 4.86 -5.12 | 4.73 - 5.31 |
| Nadir CD4+ T cell count | 230 | 155 - 5.12 | 70 - 290 |
| CD4+ T cell count pre-ART initiation | 370 | 325 - 435 | 300 - 650 |
| CD8+ T cell count pre-ART initiation | 1850 | 1360 - 2370 | 1120 - 2890 |
| CD4/CD8 ratio pre-ART initiation | 0.24 | 0.14 - 0.33 | 0.12 - 0.38 |
| CD4+ T cell count before DC vaccination (T0) | 650 | 575 - 715 | 510 - 940 |
| CD8+ T cell count before DC vaccination (T0) | 610 | 470 - 835 | 400 - 970 |
| CD4/CD8 ratio baseline (T0) | 1.21 | 0.73 - 1.39 | 0.53 - 1.48 |
| **Analytical ART interruption** | | | |
| Time to viral rebound >50 c/mL (weeks) | 3.0 | 2.3 - 3.65 | 2.0 - 6.3 |
| Time to viral rebound >500 c/mL (weeks) | 3.3 | 2.65 - 5.15 | 2.0 - 6.3 |
| Time to viral rebound >1000 c/mL (weeks) | 3.3 | 2.65 - 5.15 | 2.0 - 8.3 |
| Time to viral rebound >50000 c/mL (weeks) | 6.3 | 3.15 -20.15 | 2.0 - 40.0 |
| Lowest CD4+ T cell count | 250 | 175 - 285 | 150 - 290 |
| Peak CD8+ T cell count | 1370 | 1195 - 2040 | 1060 - 4400 |
| last CD4+ T cell count before ART resumption (T4) | 250 | 180 - 300 | 150 - 350 |
| Last CD8+ T cell count before ART resumption (T4) | 820 | 685 - 1090 | 580 - 1650 |
| CD4/CD8 ratio before ART resumption (T4) | 0.25 | 0.20 - 0.39 | 0.17 - 0.43 |
| Weeks off ART | 50 | 28.5 - 96.0 | 28 - 116 |
| **ART resumption** | | | |
| Time to viral resupression <50 c/mL (months) | 8.5 | 4.0 - 16.0 | 3.0 - 18.0 |
| Time on ART post resumption, early (years); T5 | 2.3 | 2.2 - 3.2 | 2.0 - 3.2 |
| CD4+ T cell count (T5) | 620 | 525 - 663 | 490 - 710 |
| CD8+ T cell count (T5) | 975 | 617.5 - 1098 | 520 - 1510 |
| CD4/CD8 ratio (T5) | 0.65 | 0.62 - 0.92 | 0.38 - 1.0 |
| Time on ART post resumption, late (years); T6 | 12.4 | 11.9 - 13.05 | 11. - 13.3 |
| Time on ART undetectable (years); T6 | 10.5 | 7.8 - 11.5 | 6.2 - 12.9 |
| CD4+ T cell count (T6) | 590 | 490 - 780 | 430 - 800 |
| CD8+ T cell count (T6) | 650 | 520 - 770 | 460 - 2240 |
| CD4/CD8 ratio (T6) | 1.02 | 0.67 - 1.20 | 0.30 - 1.42 |

***Note**: Three participants with viral blips (50-400 c/mL) since viral re-suppression. **H003** – 255 c/mL (Oct 2009), 132 c/mL (April 2010), 59 c/mL (Jan 2014); **H006** (53 (Feb 2003), 57 c/mL and 228 c/mL (Nov 2013); **H008** (slow to suppress after restarting ART); **H017** – 71 c/mL (Jan 2016).*

**Table S2** Annotation Clusters

| | |
|---|---|
| CD8+ TCM | CD3+CD8+CD45RA-CCR7+CD28+CD27+ |
| CD8+ TEMRA | CD3+CD8+CD45RA+CCR7-CD28-CD27- |
| CD8+ TEM CD28-CD27- | CD3+CD8+CD45RA-CCR7-CD28-CD27- |
| CD8+ TEM CD28+CD27+ | CD3+CD8+CD45RA-CCR7-CD28+CD27+ |
| CD8+ TN | CD3+CD8+CD45RA+CCR7+CD28+CD27+ |
| CD4+ TCM | CD3+CD4+CD45RA-CCR7+CD28+CD27+ |
| CD4+ TN | CD3+CD4+CD45RA+CCR7+CD28+CD27+ |
| CD4+ TEM CD28+CD27- | CD3+CD4+CD45RA-CCR7-CD28-CD27- |
| CD4+ TEM CD28-CD27- | CD3+CD4+CD45RA-CCR7-CD28-CD27- |
| T regs | CD3+CD4+CD25high |
| gdTCR CD45RA+ | CD3+gdTCR+CD45RA+ |
| gdTCR CD45RA- | CD3+gdTCR+CD45RA- |
| DN T cells | CD3+CD4-CD8- |
| DN T cells 2 | CD3+CD4-CD8- |
| NKT cells | CD3+CD8+CD56+ |
| NK CD56+CD16+CD57low | CD3-CD16+CD56+CD57- |
| NK CD56+CD16+ CD57high | CD3-CD16+CD56+CD57+ |
| NK CD56+CD16- | CD3-CD56+CD16- |
| CD56dimCD16+ CD24+CXCR3+ | CD3-CD16+CD56dim CD24+CXCR3+ |
| IgG+ B cells | CD19+CD20+HLA-DR+CD24+CD38-CD39+IgG+ |
| IgM+ IgD+ CD38+ B cells | CD19+CD20+IgM+IgD+IgG-CD38+ |
| IgM+ IgD+ CD38- B cells | CD19+CD20+IgM+IgD+IgG-CD38- |
| IgM+ IgD+ CD38- B cells 2 | CD19+CD20+IgM+IgD+IgG-CD38- |
| Plasmablasts | CD19lowCD20low HLA-DR+CD27+CD38+CD39+ |
| Classical Monocytes | HLA-DR+CD11b+CD14+CD16- |
| Classical Monocytes 2 | HLA-DR+CD11b+CD14+CD16- |
| Classical Monocytes 3 | HLA-DR+CD11b+CD14+CD16- |
| Intermediate Monocytes | HLA-DR+CD11b+CD14+CD16+ |
| Intermediate Monocytes 2 | HLA-DR+CD11b+CD14+CD16+ |
| CD11c+CD16+ DCs | HLA-DR+CD11c+CD16+ |
| CD11c+CD1c+ DCs | HLA-DR+CD11c+CD1c+ |
| pDCs | HLA-DR+CD11c-CD123+ |
| DR- Basophils | HLA-DR-CD123+CD11c-CD56-CD16- |
| MAIT cells | CD3+CD8+CD27+CD28+CD161highCCR5+CCR6+ |
